# EPIGENETIC AGE ACCELERATION, LUNG FUNCTION AND COPD ACROSS THE LIFESPAN

**DOI:** 10.1101/2025.10.27.25338710

**Authors:** Sandra Casas-Recasens, Judith M Vonk, Tigist D. Adane, Julieta Viglino, Alvar Agusti, Maarten van Den Berge, Rosa Faner, Maaike De Vries

## Abstract

**Rationale:** Chronic Obstructive Pulmonary Disease (COPD) is considered an aging-associated disease but can have its origin early in life. Chronological age is an imprecise measure of biological aging. To accurately measure biological age and age acceleration, different generations of epigenetic clocks have been developed.

**Objective:** To evaluate the role of epigenetic age acceleration in COPD across adulthood, COPD severity and smoking status and in adults younger than 50 years.

**Methods:** First-, second- and third-generation epigenetic clocks were used to estimate biological aging, telomere length and pace of aging in two cohorts (Lifelines COPD&Control DNA cohort and a Spanish COPD cohort). The association between age acceleration and FEV_1_%pred, FEV_1_/FVC and COPD was evaluated. In those clocks with significant associations, the effect of COPD severity, smoking status as well as the mediating role of age acceleration on the association between smoking and lung function levels was assessed. Finally, we investigated if these associations were already present in early adulthood.

**Results:** Age acceleration estimated by second- and third-, but not first-, generation clocks was associated with FEV_1_%pred, FEV_1_/FVC and COPD in both cohorts. Associations were strongest in severe COPD cases and current smokers and age acceleration mediated the association between smoking and lung function levels. Importantly, these associations were already present in early adulthood.

**Conclusion:** Epigenetic clock-derived age acceleration relates to lower lung function and COPD and mediated smoking-related lung impairment. As these associations already exist in early adulthood, age acceleration holds potential as tool to identify smokers susceptible to COPD at early age, potentially enabling preventive strategies.

## INTRODUCTION

Chronic obstructive pulmonary disease (COPD) is a heterogeneous airway condition, characterized by airflow limitation, chronic inflammation, loss of alveolar-capillary units, and often progressive decline in lung function. Different COPD etiotypes have been described, but cigarette smoke and aging are still the two major risk factors(1).

Aging is a natural process associated with chronological age, characterized by a progressive functional decline at the molecular, cellular, tissue, and organismal levels(2). Epigenetic alterations, such as DNA methylation, occur in all tissues as a function of aging(3). Changes in DNA methylation occur before and after birth and capture external environmental influences (i.e. exposome), such as diet, trauma, exercise, and other lifestyle factors that affect aging, disease incidence and progression(3). In monozygotic twins, differences in exposome pressure have been associated with a different pace of aging and a divergent DNA methylation profile, suggesting that chronological aging can be decoupled from biological aging(4).

To measure the chronological and biological age decoupling, several DNA methylation clocks have been developed. These clocks use mathematical algorithms to select sets of DNA methylation markers (i.e. CpGs) that estimate biological age. First generation clocks, such as the “Hannum” clock (5) and the “Horvath” clock (6) capture changes in chronological age, partly driven by age-related changes in blood cell composition. To more directly investigate biological age, a second generation of clocks has been trained on age and disease related phenotypes in combination with chronological age. Epigenetic clocks such as the "PhenoAge" clock(7), or "GrimAge" clock(8), which incorporate age-related biochemical measures lead to stronger predictions of both lifespan and health span than first generation clocks. Finally, a third generation clock, the “DunedinPACE” clock(9), integrates longitudinal measurements to determine the deterioration of system integrity over time. Additionally, the DNA methylation-based estimator of telomere length (DNAmTL) according to Horvath’s methods can estimate replicative senescence(10).

Since their introduction, epigenetic clocks have been used to explore the associations between age acceleration and a variety of outcomes and diseases. For COPD, associations between accelerated aging estimated with both first- and second-generation epigenetic clocks and mortality have been shown(11). Furthermore, the GrimAge epigenetic clock was associated with COPD in people living with human immunodeficiency virus (12) and for all three generations of epigenetic clocks as well as for methylation-based telomere length associations with self-reported COPD and emphysema have been shown(13-16). However, the association between accelerated aging and the full range of lung function impairment and COPD across the lifespan is not known.

In the current study, to establish if and to what extent accelerated aging may contribute to the development and progression of COPD, we systematically explored the association between accelerated aging across the spectrum of the different epigenetic clocks and airflow impairment, airway obstruction and presence and severity of COPD. In addition, we investigated the effect of smoking on accelerated aging and whether signs of accelerated aging can already be detected earlier in life before the onset of clinically overt COPD.

## METHODS

Full methods are described in the online supplement.

### Study population

We included in this analysis data from two independent studies: the Lifelines COPD & Controls DNA methylation study (LL COPD&C) from the Lifelines population-based cohort study (17, 18) and the clinical COPD study from Spain (COPD-Sp, i.e. BIOMEPOC and EarlyCOPD cohorts)(19-21).

### Measurements

#### Forced spirometry

In LL COPD&C, pre-bronchodilator spirometry was performed according to the ATS/ERS guidelines. In COPD-sp, forced spirometry and bronchodilator test upon 400 mg salbutamol was conducted in accordance with GOLD recommendations. In both cohorts, following the GOLD recommendations, COPD was defined by a ratio between forced expiratory flow within 1 second (FEV_1_) and forced vital capacity (FVC) below 0.7(22).

#### Genome wide DNA methylation

In LL COPD&C, DNA methylation was measured with the Infinium HumanMethylation450 Beadchip (Illumina). In COPD-Sp, the Infinium MethylationEPIC Beadchip was used (Illumina)(23).

### Epigenetic age

We estimated: (1) biological age with the first- and second-generation Horvath, Hannum, SkinHorvath, Best Linear Unbiased Prediction (BLUP), Elastic Net (EN), PhenoAge and GrimAge epigenetic clocks(5-8, 24, 25); (2) pace of aging with the third-generation DunedinPACE epigenetic clock(9); and (3) telomere length with the DNA methylation-based estimator of telomere length (DNAmTL)(10).

### Age acceleration

Age acceleration was calculated by regressing DNA methylation-derived biological age on chronological age and taking the residual value. For DunedinPACE, the estimated pace of aging was used as age acceleration. To be able to compare the effect sizes of the clocks, age acceleration was standardized by dividing the calculated age acceleration by their respective standard deviation. In the analyses, the effect size is expressed as units of standard deviation.

### Statistical analysis

For all analyses, a p-value of 0.05 was considered significant.

#### Correlation between chronological and biological age

The linear correlation between chronological age and the DNA methylation-derived biological age was assessed using linear regression with biological age as outcome adjusted for sex and smoking status.

#### Association between accelerated aging and lung function levels and (severity of) COPD

To assess the association between accelerated aging and FEV_1_%predicted and FEV_1_/FVC, we performed linear regression analysis. The association between accelerated aging and COPD and severity of COPD was assessed using logistic and multinomial logistic regression analysis, respectively. For severity of COPD, mild COPD was used as reference. All models were adjusted for sex and smoking status.

The aforementioned analyses were stratified by smoking status (never/current in LL COPD&C; former/current in COPD-Sp) and we examined the interactions between age acceleration and smoking for all epigenetic clocks that showed a significant association with at least one lung function outcome.

#### Mediation analysis

To investigate if the association between smoking and lung function was mediated by age acceleration, we performed a causal mediation analysis using the r package mediation. For all epigenetic clocks, the average causal mediation effect (ACME), average direct effect (ADE), total effect and proportion mediated were calculated.

#### Young individuals

In LL COPD&C, we additionally performed the analyses described above in individuals younger than 50 years of age.

## RESULTS

### Participant characteristics

Table 1 shows the main clinical characteristics of both LL COPD&C and COPD-Sp cohorts. The median age was lower and lung function levels higher in LL COPD&C compared to COPD-Sp. By using both cohorts, we cover the full spectrum of adult lifespan (from 37 to 88 years of age) and airflow obstruction severity (mild to very severe).

**Table 1.**
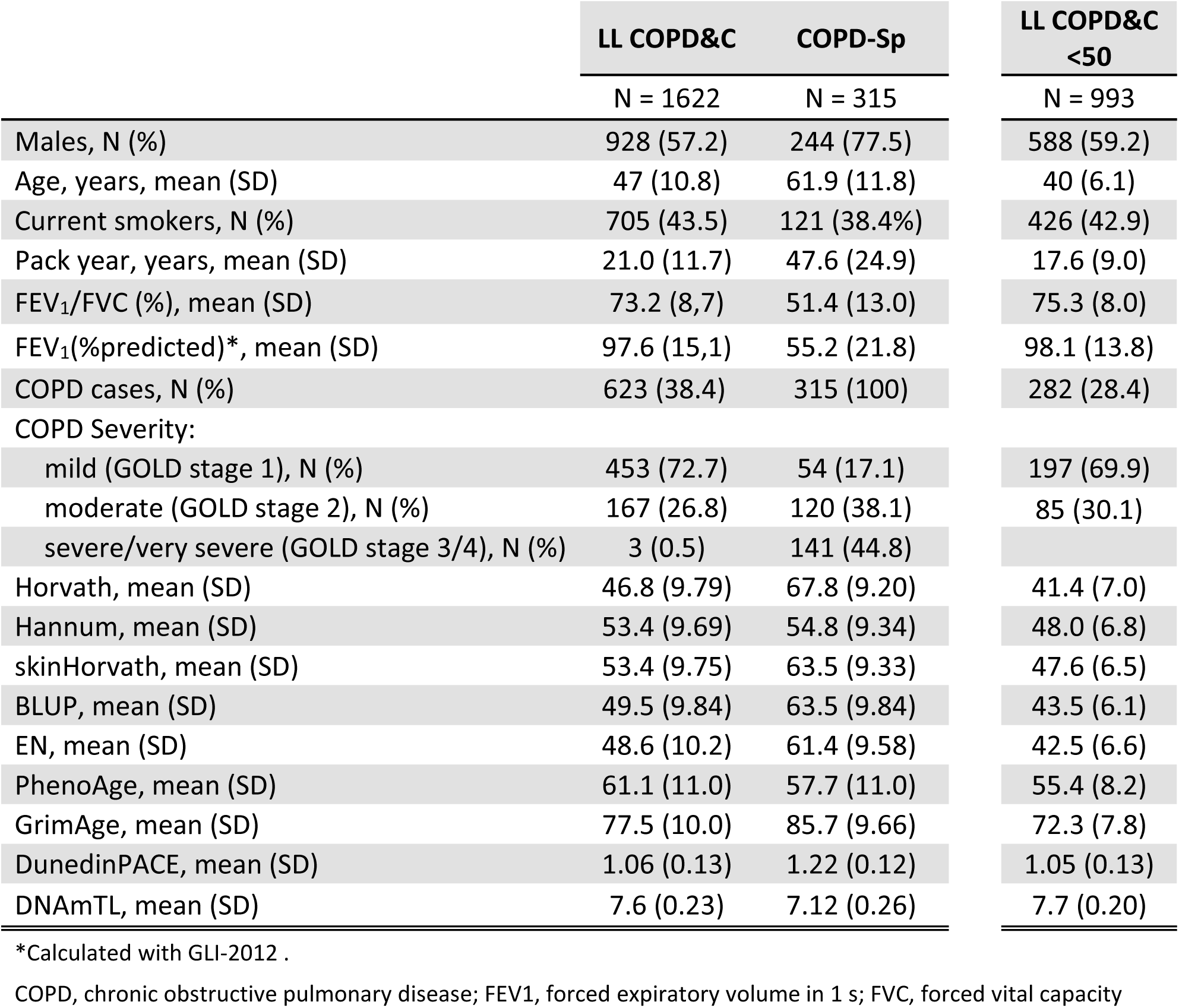
Clinical characteristics.

### Epigenetic age is significantly associated with chronological age

Epigenetic age estimated by first-generation clocks outperformed the other clocks when investigating the association with chronological age in both the LL COPD&C and COPD-Sp cohort (Figure E1 and Table E1A). The second-generation epigenetic clocks, represented by PhenoAge and GrimAge, were also highly associated with chronological age in both cohorts, but especially for GrimAge, the estimated epigenetic age was much higher than the chronological age. DunedinPACE, the third-generation clock, was weakly associated with chronological age. Lower telomere length estimated with the DNAm-TL epigenetic clock was associated with higher chronological age.

### Age acceleration is associated with lung function levels and (severity of) COPD

We tested the association between age acceleration and FEV_1_%predicted, FEV_1_/FVC and COPD (in LL COPD&C only) for all epigenetic clocks (Table E2). In Figure 1, the results for the clocks that showed at least one significant association with one of the three outcomes are presented. For the first-generation clocks, no significant associations were found. For the second-generation clocks, PhenoAge- and GrimAge-estimated accelerated aging was significantly associated with lower FEV_1_%predicted and FEV_1_/FVC in both cohorts, except for PhenoAge on FEV_1_/FVC in LL COPD&C (p=0.11). In addition, GrimAge but not PhenoAge (p=0.079) was associated with COPD in LL COPD&C. Representing the third-generation clocks, DunedinPACE was associated with lower FEV_1_%predicted and FEV_1_/FVC in both cohorts together with higher odds of COPD in LL COPD&C. For telomere length, we observed a significant association between longer telomeres and higher FEV_1_%predicted and FEV_1_/FVC in COPD-Sp.

**Figure 1:**
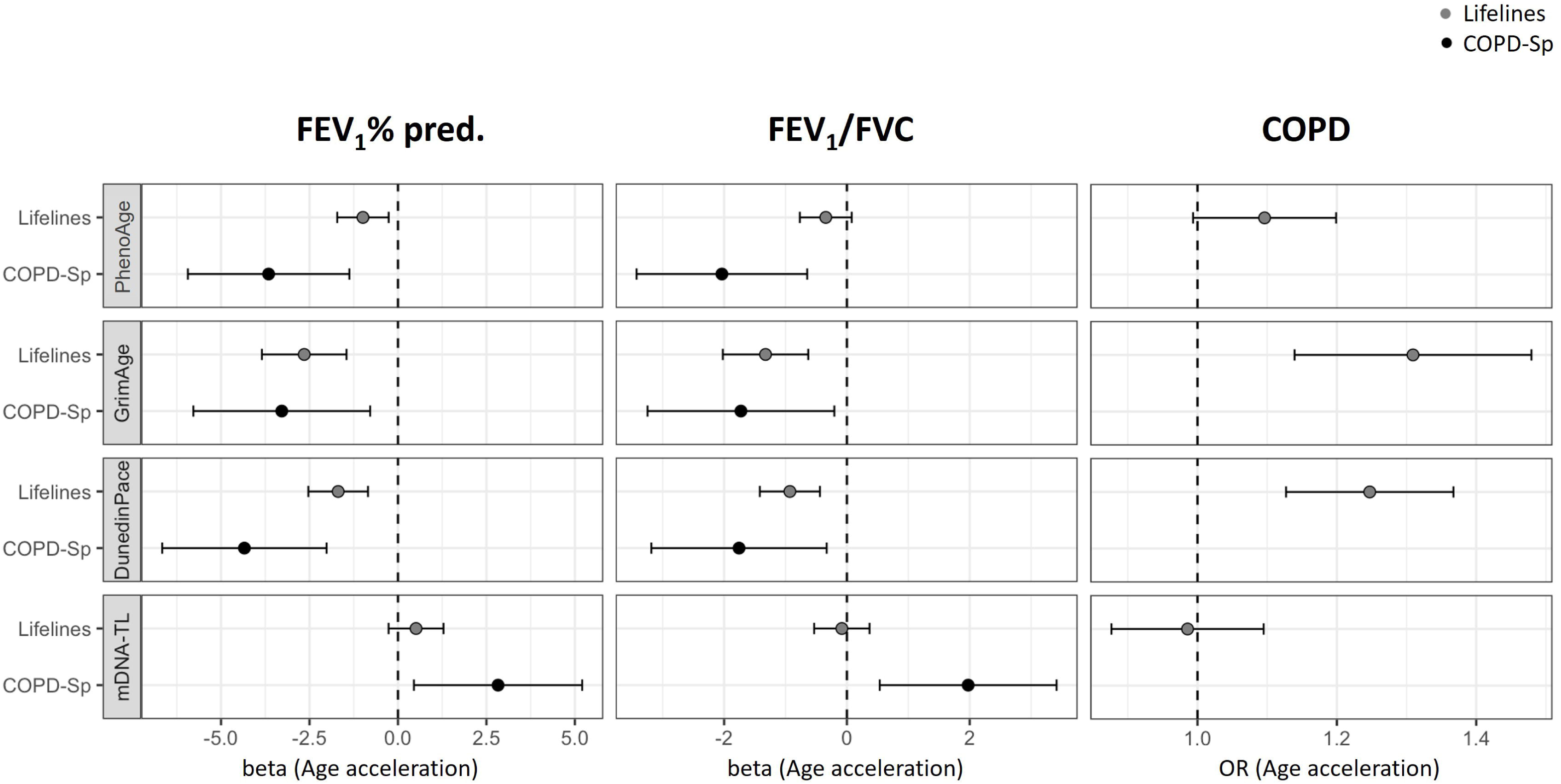
Associations between age acceleration and FEV_1_%predicted, FEV_1_/FVC, and COPD (in LL COPD&C only) for the clocks presenting at least one significant association with any of the outcomes. Grey circles for LL COPD&C cohort and black circles for COPD-Sp cohort. Forest plots of the linear regression models on FEV_1_%predicted and FEV_1_/FVC presenting the point estimates and 95% CI (whiskers), as units of standard deviation of age-acceleration or the logistic regression model on COPD presenting the OR and 95% CI (whiskers). All models adjusted for sex and smoking. Standard deviation for PhenoAge: 5.72 (LL COPD&C) and 6.54 (COPD-Sp); GrimAge: 5.78 (LL COPD&C) and 3.91 (COPD-Sp); DunedinPACE: 0.13 (LL COPD&C) and 0.12 (COPD-Sp); mDNA-TL: 0.16 (LL COPD&C) and 0.18 (COPD-Sp).

Additionally, we investigated if age acceleration was associated with COPD severity, as reflected by GOLD stages, using mild COPD (GOLD1) as reference. Off note, in LL COPD&C, the three participants with severe (GOLD3) COPD were analyzed together with those with moderate (GOLD2) COPD. As shown in Table 2, DunedinPACE was associated with moderate COPD in LL COPD&C and with both moderate and severe-very severe (GOLD3-4) COPD in COPD-Sp. Additionally in COPD-Sp, the second-generation epigenetic clocks PhenoAge and GrimAge were both associated with severe-very severe COPD.

**Table 2.**
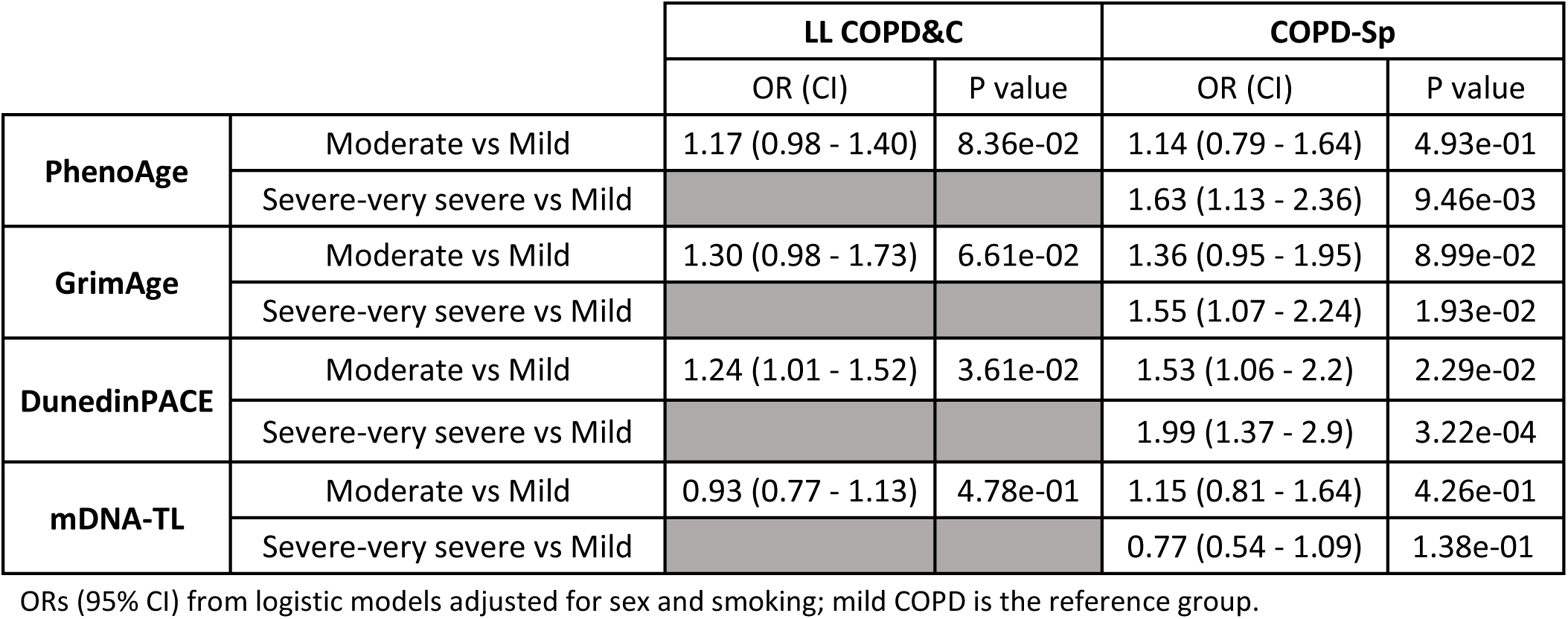
Model coefficients for age acceleration in the association with COPD severities for selected clocks.

|  |  | LL COPD&C |  | COPD-Sp |  |
| --- | --- | --- | --- | --- | --- |
|  |  | OR (CI) | P value | OR (CI) | P value |
| **PhenoAge** | Moderate vs Mild | 1.17 (0.98 - 1.40) | 8.36e-02 | 1.14 (0.79 - 1.64) | 4.93e-01 |
|  | Severe-very severe vs Mild |  |  | 1.63 (1.13 - 2.36) | 9.46e-03 |
| **GrimAge** | Moderate vs Mild | 1.30 (0.98 - 1.73) | 6.61e-02 | 1.36 (0.95 - 1.95) | 8.99e-02 |
|  | Severe-very severe vs Mild |  |  | 1.55 (1.07 - 2.24) | 1.93e-02 |
| **DunedinPACE** | Moderate vs Mild | 1.24 (1.01 - 1.52) | 3.61e-02 | 1.53 (1.06 - 2.2) | 2.29e-02 |
|  | Severe-very severe vs Mild |  |  | 1.99 (1.37 - 2.9) | 3.22e-04 |
| **mDNA-TL** | Moderate vs Mild | 0.93 (0.77 - 1.13) | 4.78e-01 | 1.15 (0.81 - 1.64) | 4.26e-01 |
|  | Severe-very severe vs Mild |  |  | 0.77 (0.54 - 1.09) | 1.38e-01 |

ORs (95% CI) from logistic models adjusted for sex and smoking; mild COPD is the reference group.

### Association between age acceleration and lung function levels and COPD stratified by smoking status

Next, to determine whether the association between age acceleration and lung function levels and COPD depends on smoking status, we conducted stratified analyses (Figure 2 and Table E3). In LL COPD&C, the associations between age acceleration and all lung function outcomes are larger in current smokers compared to never smokers. These associations were significant in current smokers for the second and third generation clocks. A similar pattern was observed in COPD-Sp, where the associations were stronger in current smokers than in former smokers with significant associations primarily found in current smokers for second and third generation clocks. Importantly, interaction analyses showed that, especially in LL COPD&C, many of these differences between never/former and current smokers were significant (Figure 2).

**Figure 2:**
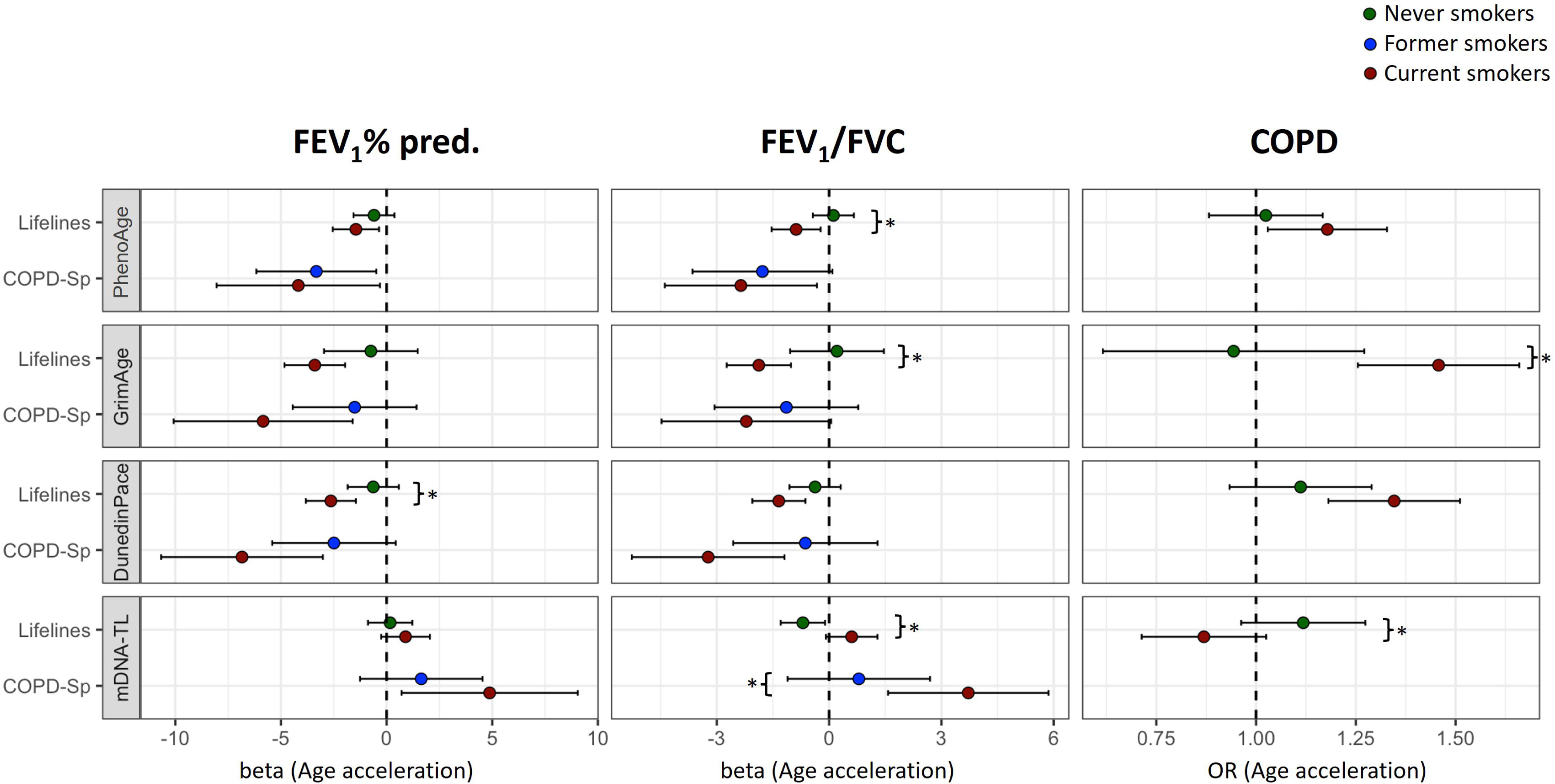
Associations between age acceleration and FEV_1_%predicted, FEV_1_/FVC, and COPD (in LL COPD&C only), stratified by smoking status (never-smoker (NS) green circles, current-smoker (CS) red circles and former-smoker (FS) blue circles), for the clocks presenting at least one significant association with any of the outcomes. Forest plots of the linear regression models on FEV_1_%predicted and FEV_1_/FVC presenting the point estimates and 95% CI (whiskers), as units of standard deviation of age-acceleration or the logistic regression model on COPD presenting the OR and 95% CI (whiskers). All models adjusted for sex. Asterix (*) indicates a significant interaction of p<0.05 between smoking and the respective age acceleration.

Smoking-stratified analysis on COPD severity (with mild COPD as reference) did not show consistent differences between current smokers and never/former smokers (Table E4).

### Age acceleration mediated the association between smoking and lung function

We tested if the association between current smoking and lung function was mediated by age acceleration. In LL COPD&C, GrimAge-derived age acceleration and DunedinPACE-derived pace of aging significantly mediated the association between smoking and both FEV_1_% predicted (64% and 28% of the total effect of smoking on FEV1%predicted was mediated by age acceleration respectively) and FEV_1_/FVC (73% and 35% mediation, respectively) (Figure 3 and Table E5). In addition, PhenoAge-derived age acceleration mediated the association between smoking and FEV_1_% predicted (5.3% mediation). In COPD-Sp, GrimAge-derived age acceleration significantly mediated the association between smoking and FEV_1_% predicted (-16% mediation) and FEV_1_/FVC (-17% mediation). However, the direction of mediation was opposite to what was observed in LL COPD&C.

**Figure 3:**
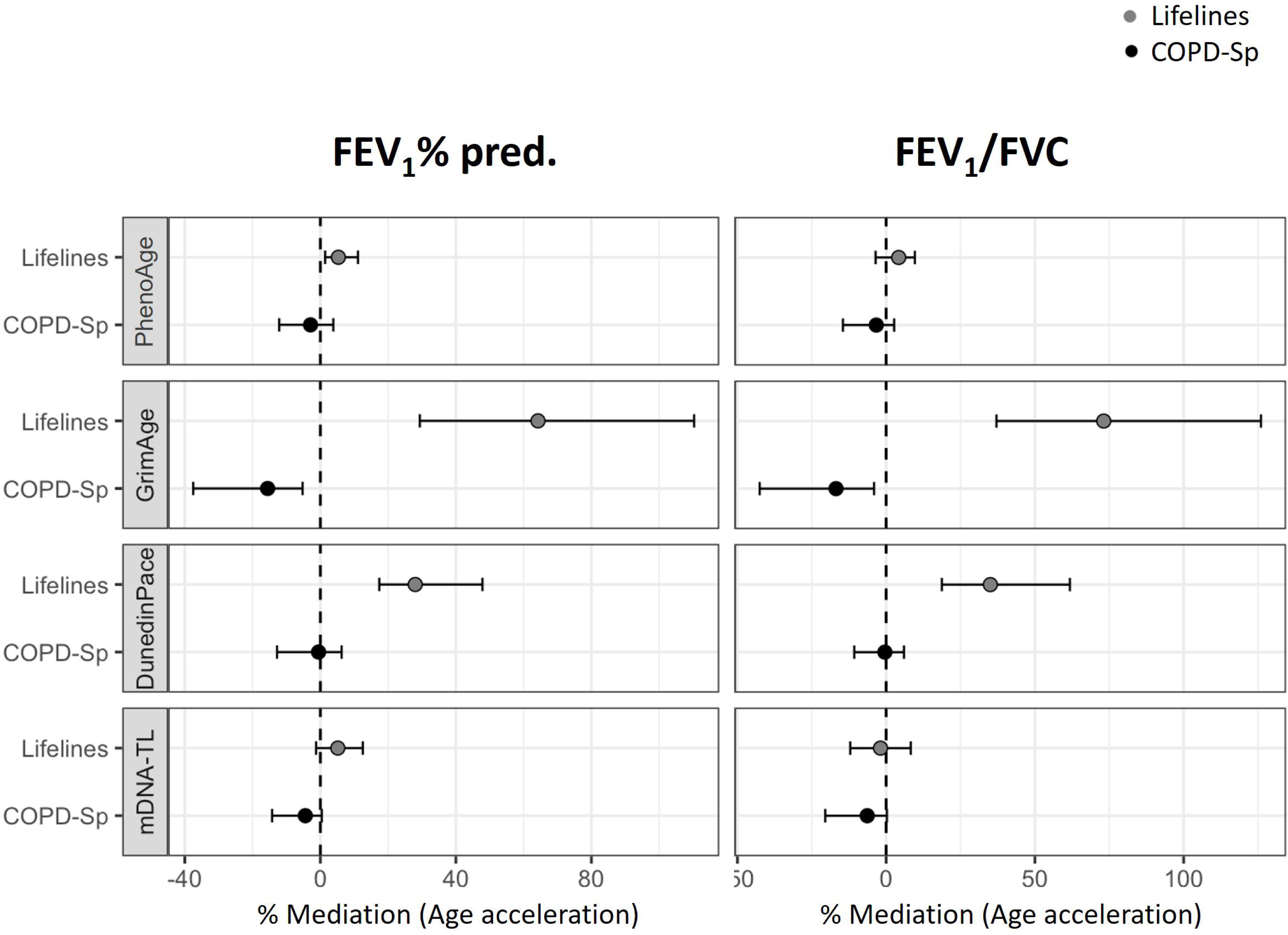
The mediation effect of age acceleration estimated with the PhenoAge, GrimAge, DundedinPACE and mDNA-TL epigenetic clock on the association between smoking and FEV_1_% predicted and FEV_1_/FVC. Grey circles for LL COPD&C cohort and black circles for COPD-Sp cohort. Forest plots indicate the percentage of mediation of age acceleration on the association between smoking and lung function.

### Age acceleration association in young individuals

Finally, we tested whether the associations between age acceleration and FEV_1_ % predicted, FEV_1_/FVC and COPD were already present in those younger than 50 years from the LL COPD&C cohort across all epigenetic clocks. In this case, PhenoAge, GrimAge and DunedinPACE, showed significant negative associations with all outcomes (Figure 4 and Table E6A). Regarding COPD severity, only GrimAge was significantly associated with moderate COPD in young individuals (Table E7A).

**Figure 4:**
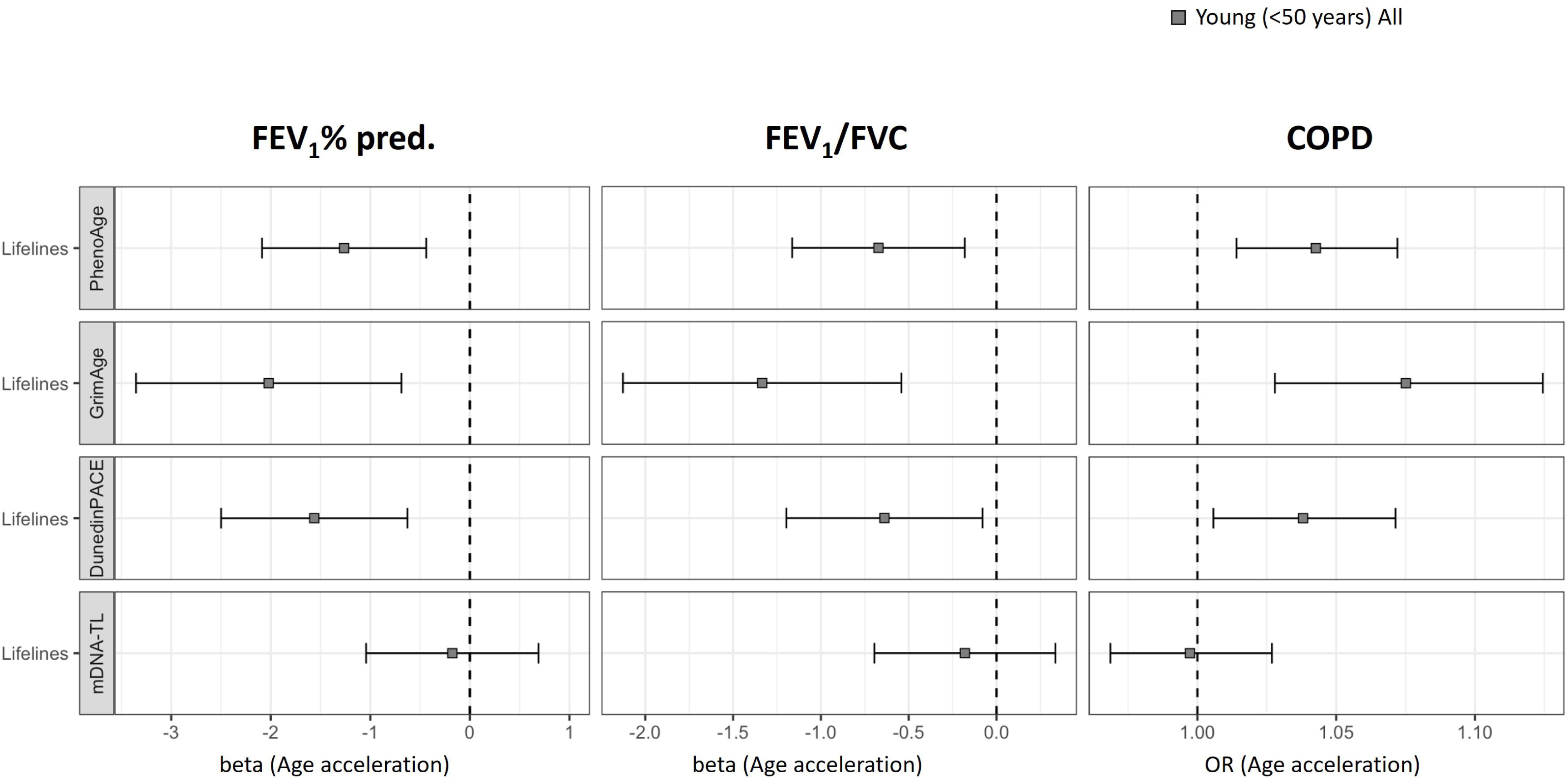
Associations between age acceleration and FEV_1_%predicted, FEV_1_/FVC, and COPD, in young individuals from LL COPD&C (<50 years old). Forest plots of the linear regression models on FEV_1_%predicted and FEV_1_/FVC presenting the point estimates and 95% CI (whiskers), as units of standard deviation of age-acceleration or the logistic regression model on COPD presenting the OR and 95% CI (whiskers). All models adjusted for sex and smoking.

Stratification by smoking status revealed that the associations between age acceleration and lung function were consistently stronger among young current smokers compared to young never smokers (Figure 5 and Table E6B). Stratifying for smoking status did not reveal significant associations of age-acceleration with moderate COPD in young individuals (Table E7B).

**Figure 5:**
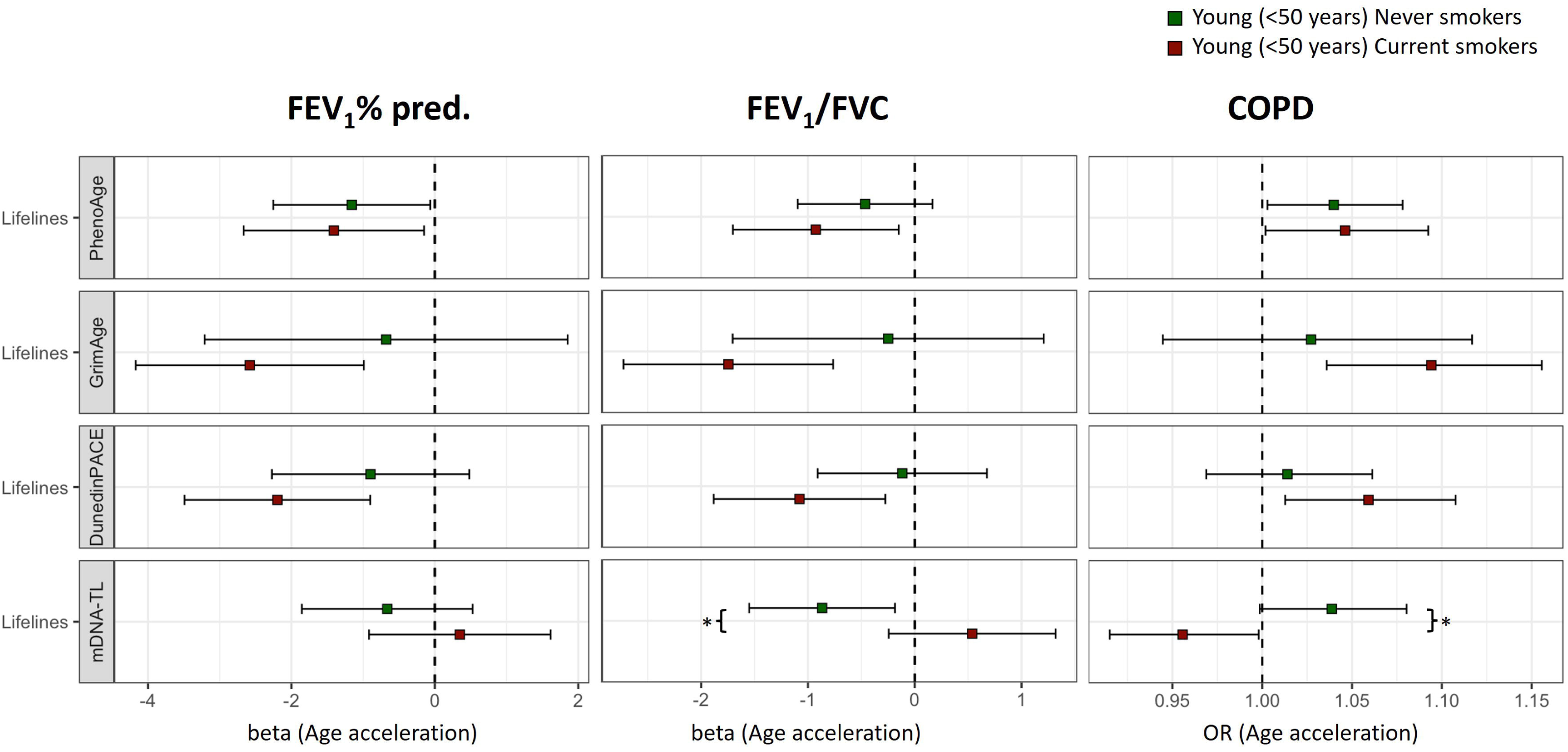
Associations between age acceleration and FEV_1_%predicted, FEV_1_/FVC, and COPD, in young individuals from LL COPD&C (<50 years old), stratified by smoking status (never-smoker (NS) green squares, current-smoker (CS) red squares), for the clocks presenting at least one significant association with any of the outcomes. Forest plots of the linear regression models on FEV_1_%predicted and FEV_1_/FVC presenting the point estimates and 95% CI (whiskers), as units of standard deviation of age-acceleration or the logistic regression model on COPD presenting the OR and 95% CI (whiskers). All models adjusted for sex.

PhenoAge-, GrimAge- and DunedinPACE-derived age acceleration significantly mediated the association between current smoking and FEV_1_ %predicted and FEV_1_/FVC in young individuals (Figure 6 and Tabel E8).

**Figure 6:**
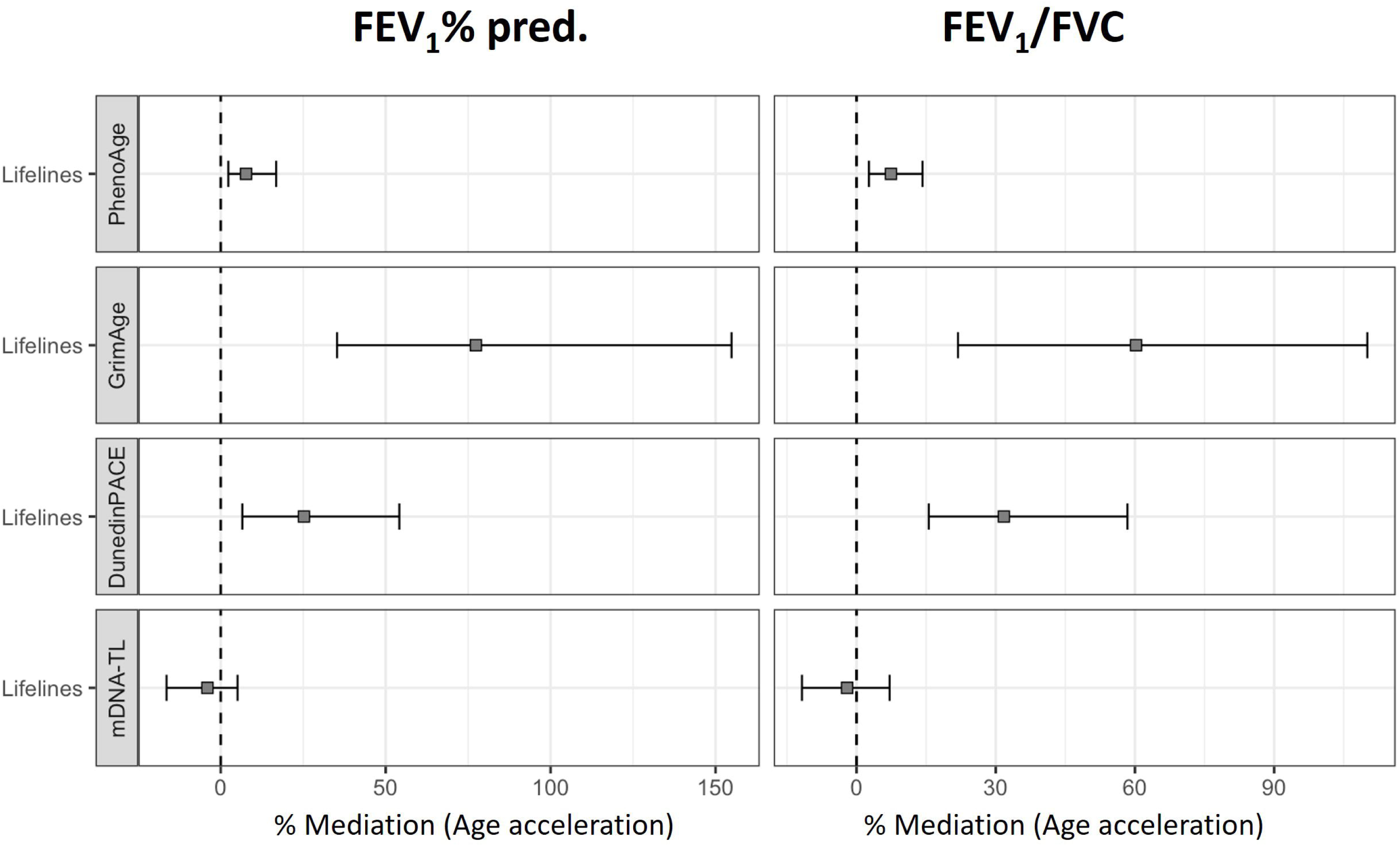
The mediation effect of age acceleration estimated with the PhenoAge, GrimAge, DundedinPACE and mDNA-TL epigenetic clock on the association between smoking and FEV_1_% predicted and FEV_1_/FVC, in young individuals from LL COPD&C (<50 years old). Forest plots indicate the percentage of mediation of age acceleration on the association between smoking and lung function.

## DISCUSSION

In this study, we investigated the association between all three generation epigenetic clocks and lung function levels and COPD, not only across adulthood and the full spectrum of airflow limitation severities, but also in adults younger than 50 years of age. The main findings were: (1) age acceleration measured by the second and third generation clocks, but not the first generation clocks, was significantly associated with the severity of airflow limitation (measured by FEV1 % predicted) and airway obstruction (measured by FEV1/FVC), and with the presence and severity of COPD. (2) Accelerated aging has a higher impact on COPD and lung function levels in the presence of smoking and partly explains the detrimental effect of smoking on lung function levels. (3) Associations with accelerated aging can already be identified in individuals younger than 50 years of age, suggesting that accelerated aging may predispose to COPD earlier in life.

We found that accelerated aging estimated with the second- and third-generation epigenetic clocks was associated with airflow impairment, airway obstruction and presence and severity of COPD, which was not found for age acceleration estimated with the first-generation epigenetic clocks. In this context, it is important to consider how the different clocks were trained(26, 27). The first-generation clocks were trained purely based on chronological age and are less sensitive to the effects of smoking and health outcomes. In contrast, health outcomes were taken into account for the training of the second-generation clocks. For DunedinPACE, the third-generation epigenetic clock, various longitudinal organ-system integrity biomarkers were used for its training. With this longitudinal approach, they are expected to offer more precise identification of trends in lung function decline and disease progression compared to the cross-sectional designs of earlier clocks. In line with the clock-specific features, it is not surprising that GrimAge, PhenoAge and DunedinPACE exhibited the strongest associations with lung function traits and COPD in our study. While poor correlations between first-generation clocks and lung function levels have previously been shown(13, 15), significant associations were found for the second-generation clocks(12, 14, 15). Specifically, GrimAge-derived age acceleration has been associated with FEV_1_, FEV_1_/FVC, maximum mid-expiratory flow and COPD(12, 14, 15), and PhenoAge-derived age acceleration with FEV_1_% predicted and FEV_1_/FVC(7). Recently, higher pace of aging as estimated with the third-generation DunedinPACE epigenetic clock was observed with self-reported COPD and emphysema(16). To mention, the association between methylation-derived telomere length and lung function traits was significant only in the COPD-Sp cohort, characterized by more severe lung impairment. This finding suggests that telomere attrition becomes detectable primarily in advanced stages of disease, maybe reflecting the cumulative impact of chronic inflammation and oxidative stress on cellular aging.

Interestingly, we show that the magnitude of the associations between age acceleration and airflow impairment, airway obstruction and the presence and severity of COPD is stronger in current smokers compared to never or former smokers. Likely, this is due to chronic and systemic inflammation caused by current smoking, known to impact both age acceleration and lung function decline. However, also the incorporation of smoking into the epigenetic clock algorithms may play a role. Specifically, the second-generation GrimAge clock directly incorporates smoking effects, rendering it the most sensitive to smoking-related changes(8). PhenoAge captures smoking effects indirectly through its influence on clinical biomarkers of aging(7), while for DunedinPACE, the impact of smoking is reflected by capturing its physiological consequences(27). The observation that the effect of smoking is only significantly different for some of the traits in LL COPD&C, but not in COPD-sp, may partly reflect the differences in cohort profiles. In LL COPD&C, a non-random subset of the population-based Lifelines cohort study with predominantly individuals with mild COPD, only never and current smokers were included. In contrast, the COPD-Sp cohort is a clinical cohort including only ever smokers with COPD across all severities of airflow limitation. This suggests that a history of smoking continues to influence epigenetic aging, as reflected by the CpG-sites used in the PhenoAge and GrimAge epigenetic clocks. Another possible explanation is that, in the COPD-Sp cohort, former smokers tend to have more severe airflow limitation, while current smokers typically have milder airflow limitation. Since both smoking history and airflow limitation severity contribute to age acceleration, their combined effects may counterbalance each other, making the interaction between smoking and age acceleration less detectable in this cohort. Nevertheless, a lack of power could also play a role here. Furthermore, our analyses suggest that current smoking leads to higher biological age, which, in turn, results in lower lung function. In other words, age acceleration partly explains the lower lung function levels that occur in smokers. In this context, it is of interest that former smokers had a lower lung function compared to current smokers in COPD-sp, leading to an opposite mediation effect for GrimAge-derived age acceleration. Although this may seem contradictory, it can be explained by the fact that those with worse COPD and more severe disease symptoms were more likely to quit smoking. Notably, among moderate COPD patients in COPD-sp, a higher percentage are current smokers, and they exhibit similar chronological age and cumulative smoking exposure to those with severe/very severe COPD. This could suggest that individuals with severe obstruction may represent a subgroup that is more vulnerable to environmental exposures contributing to COPD and/or possess a genetic or epigenetic predisposition to accelerated biological aging beyond the effects of smoking.

Beyond the smoking-related effects, we also showed that the association between accelerated aging and airflow impairment, airway obstruction and presence of COPD can already be observed in individuals younger than 50 years of age. To some extent, this supports the finding by Perez-Garcia *et al* showing a higher HannumAge for individuals reporting onset of COPD before age of 40 compared to those with late onset(16). Together, this suggests that age acceleration is already present in early life, potentially even before the onset of overt COPD, especially in those who smoke. This observation is important because it suggests that assessment of accelerated aging can be useful as a tool to identify smokers susceptible to future lung function loss before development of overt COPD, thereby enabling early interventions or preventive strategies.

Among the strengths of this study are that it is one of the first to evaluate age acceleration using multiple epigenetic clocks and examine their associations with lung function parameters, COPD and COPD severity, stratified by smoking and in a subgroup of young individuals. The analysis was conducted across two distinct cohorts: a population-based cohort that includes healthy controls and subjects with mild COPD and a COPD cohort comprising patients with varying severity levels. This dual-cohort approach provides both a comprehensive overview and specific insights into the relationship between lung function levels, disease progression, and accelerated aging, enhancing our understanding of how aging dynamics intersect with respiratory health. However, we acknowledge some potential limitations. Firstly, epigenetic age was derived from blood samples, which may not fully capture organ-specific aging processes. Nevertheless, previous studies have demonstrated that epigenetic aging measured in blood correlates with lung-related diseases, such as lung cancer(28). Secondly, as mentioned in the methods, in LL COPD&C only pre-bronchodilator spirometry was available, whereas COPD-SP spirometry measurements were post-bronchodilator. Post-bronchodilator values better reflect structural lung aging, while pre-bronchodilator values also capture reversible obstruction, increasing variability and potentially weakening associations. However, due to large sample size of the Lifelines cohort study, post-bronchodilator spirometry was not feasible. Lastly,the cross-sectional nature of the study limits our ability to infer temporal causality. Longitudinal studies are needed to validate these findings and determine whether accelerated epigenetic aging contributes causally to lung function decline or is merely a consequence of disease progression.

In conclusion, we show that accelerated aging is associated with airflow impairment, airway obstruction and the presence and severity of COPD, especially in those who smoke. Importantly, accelerated aging can already be identified in young individuals with COPD. These findings suggest that aging-related biological changes emerge early in the disease course and its continuous effect may contribute to the progression of the disease at a relatively early age. Therefore, accelerated aging could help to identify COPD in at-risk populations, allowing earlier intervention or preventive strategies.

## Supporting information

Full methods

## Data Availability

All data produced in the present study ara available upon reasonable request by the authors

## ACKNOWLEDGEMENTS

Authors thank all participants for their willingness to contribute to medical research and all field personnel for their work and dedication, and all the participants and researchers of the BIOMEPOC and EarlyCOPD cohorts.

