## Supplementary material for "EPIGENETIC AGE ACCELERATION, LUNG FUNCTION AND COPD ACROSS THE LIFESPAN": Full methods

#### Study population

LL COPD&C is a non-random selection from the Lifelines cohort study based on airway obstruction ($FEV_1$/FVC and LLN < 0.7), self-reported smoking status (never smokers versus current smokers with at least 5 pack years) and occupational exposures. Therefore, the percentages of subjects with airway obstruction should not be interpreted as population prevalence. Lifelines was approved by the medical ethics committee of the UMCG (UMCG METc 2007/152) and all participants have provided signed written informed consent

The COPD-sp is a study including COPD patients from different severity stages, visited in primary care, secondary or tertiary referral hospitals in Spain. COPD-sp was approved by the local university hospital ethical committee (HCB-2018/135) and all participants have provided signed written informed consent.

#### Measurements

Lung function

In LL COPD&C, pre-bronchodilator spirometry was performed with a Welch Allyn Version 1.6.0.489, PC-based Spiroperfect with CA Workstation software according to ATS/ERS guidelines. Results and technical quality were evaluated by well-trained assistants. Results that were difficult to interpret were re-evaluated by a lung physician.

In COPD-Sp forced spirometry and bronchodilator test were standardized in accordance with the Spanish Society of Pneumology and Thoracic Surgery (SEPAR) manual of procedures for assessing lung function. The bronchodilator test was performed with the administration of 400 mg salbutamol using a spacer, in accordance with GOLD recommendations.

In both cohorts, COPD was defined by their respective spirometry as the ratio between forced expiratory flow within 1 second and forced vital capacity below 70 percent.

Genome wide DNA methylation

In LL COPD&C, DNA was isolated from whole blood at the baseline visit and 500 ng DNA was bisulfite converted using the EZ-96 DNA methylation kit (Zymo research Corporation, CA, USA). DNA methylation was measured with the Illumina Infinium Human Methylation 450K array (Illumina Inc). Quality control was performed using the Minfi package in R and the data was normalized using DASEN implemented in the watermelon package in R. Probes on sex chromosomes, probes measuring SNPs or probes with a SNP in the CpG itself or the single base extension site were removed [1]. In COPD-sp, DNA was extracted from whole blood using the QIAmp DNA Mini kit (Qiagen, Valencia, US). Its quality and quantity were assessed using Qubit fluorometer (Thermofisher). Bisulfite conversion of DNA samples was done with the EZ-96 DNA Methylation Kit (Zymo research, CA, US) and DNA methylation was measure using Infinium MethylationEPIC Beadchip [2] (Illumina). The 'ChAMP' pipeline was used for quality control, normalization and covariates/batch adjustment and differential methylation analysis of the EPIC raw data [3]. Probes containing a CpG overlying a SNP or located on sex chromosomes were removed. The type-2 probe correction BMIQ was used, and batch effects were adjusted using Combat function.

**Epigenetic age**

For the first generation of epigenetic clocks, we estimated epigenetic age with the Horvath, Hannum, SkinHorvath, Best Linear Unbiased Prediction (BLUP) and Elastic Net (EN) epigenetic clocks. The Horvath clock was designed to predict chronological age in individuals along the whole lifespan, includes 353 CpGs and was constructed across multiple tissues [4]. The Hannum epigenetic clock comprises 71 CpGs selected from the Illumina 450k array that strongly capture associations with chronological age [5]. SkinHorvath aims to predict chronological age more accurately and is additionally trained on *ex vivo* experiments and blood samples and is based on 391 CpGs [6]. As indicated by their names, BLUP (319,607 CpGs) and EN (515 CpGs) were developed using best linear unbiased prediction and elastic net, respectively, to create a clock with the highest accuracy on chronological age prediction [7].

For the second generation of epigenetic clocks, we used the PhenoAge and GrimAge epigenetic clocks, which additionally used clinical biomarkers and environmental factors to predict biological age and mortality risk. PhenoAge, developed by Levine *et al* in 2018, is generated based on a weighted average of 9 clinical characteristics and chronological age that were associated with DNA methylation levels to select 513 CpGs [8]. The GrimAge clock was developed in 2019 and is composed of seven DNA methylation-based estimators of plasma protein levels and smoking pack-years [9]. This clock (1030 CpGs) can predict lifespan but also provides information on risk of age-related conditions.

As third generation clock, we used DunedinPACE to estimate the pace of aging. DunedinPACE is developed using the rate of change in 19 blood-chemistry and organsystem-function biomarkers at four successive time points [10]. DunedinPACE includes 173 CpGs. Finally, we used the DNA methylation-based estimator of telomere length (DNAmTL) according to Horvath's methods to estimate replicative senescence [11].

Except for GrimAge, the r package methylclock [12] and the r package DunedinPACE [10] were used to estimate the different epigenetic ages in both cohorts. For GrimAge [9], access was granted by the developing team and the script was made available to use locally for LL COPD&C only; for COPD-Sp the online DNA Methylation Age Calculator [4] was used.

### SUPPLEMENTARY FIGURES

**Figure E1**: Scatter plots showing the Pearson correlation (R) between epigenetic age (calculated using all clocks) and chronological age in the LL COPD&C and COPD-Sp cohorts. Correlation lines and R values are shown for each cohort.
